# Acupressure alleviates pain and clinical symptoms in patients with sickle cell disease

**DOI:** 10.1101/2025.04.29.25326671

**Authors:** Lina Houran, Andrew Q. Pucka, Mio Jiang, Ziyue Liu, Andrew RW O’Brien, Steven E Harte, Richard E Harris, Zahra Pakbaz, Ying Wang

## Abstract

Sickle Cell Disease (SCD) is a genetic hematological disorder characterized by chronic pain, recurrent vaso-occlusive crises featuring extremely painful episodes, and other co-morbidities. Management of pain in SCD relies on persistent and/or high doses of opioids as the mainstay, which is known to be associated with significant risks of side effects, tolerance, opioid-induced hyperalgesia, and/or lower quality of life (QoL). Moreover, many SCD individuals self-manage their pain at home, which further increases the potential risks of abuse and side effects of medications. Preliminary results from our ongoing RCT demonstrate the feasibility and efficacy of self-administered acupressure as a new telehealth approach in SCD for managing pain and co-morbidities (ClinicalTrials.gov Identifier: NCT06511453). Twenty-three participants with SCD receiving self-administered acupressure every other day for five weeks plus usual care showed progressive (weekly observation during the treatment period) and sustained (monthly observation up to 6 months following the last treatment) improvements in pain, co-morbidities (e.g., physical dysfunction, fatigue, sleep disturbance, and emotional distress) and QoL as compared to the 14 participants with SCD receiving usual care alone. No significant improvements were observed throughout the observational period in individuals with usual care only. No side effects were reported from the participants receiving self-administered acupressure. These results highlight acupressure’s potential as a safe and effective telehealth approach for managing pain in SCD. A larger sample size from continuous enrollment is needed to validate these findings.

## 1. Introduction

Sickle Cell Disease (SCD) is an inherited blood disorder characterized by chronic pain and recurrent, severely painful, vaso-occlusive crises (VOCs). Current pain management strategies often rely on high-dose opioid therapy, which carries significant risks of tolerance, opioid-induced hyperalgesia, and diminished quality of life (QoL), and are otherwise limited in available treatment approaches. Acupressure is a non-invasive integrative therapy that involves applying pressure to specific acupoints, using fingers or other probes. Acupressure has demonstrated efficacy in managing various pain-related conditions,^1^ including improving pain and QoL associated with cancer,^2^ dysmenorrhea^3^ and labor.^4^ More importantly, recent studies also showed that self-administered acupressure improves pain and co-morbidities in individuals with lower back pain,^5^ and breast cancer survivors.^6^ It is known that many SCD individuals self-manage their pain^7,8^ and VOCs^9^ at home, largely owing to barriers in the healthcare system. Consequently, there is an urgent need to develop safe, effective, and cost-efficient telehealth strategies for SCD pain management. This paper reports some preliminary findings from our ongoing RCT (ClinicalTrials.gov Identifier: NCT06511453) evaluating the feasibility and effectiveness of self-administered acupressure for the management of SCD pain and co-morbidities.

## 2. Methods

### Study participants

A total of forty-five participants with SCD, aged 13-64 years (one subject aged 13 was included in the analysis, a minor deviation of inclusion criteria), were screened and enrolled from two sites (Indiana University, Indianapolis, IN; University of California, Irvine, CA). The study was approved by the Institutional Review Board at Indiana University, and all participants gave electronic written informed consent. Demographic information is shown in **Table 1**. The study started with enrollment of n=12 participants assigned to the acupressure group for assessment of safety and feasibility. The subsequent 33 participants were randomized to receive either 5 weeks of self-administered acupressure in conjunction with usual care or usual care alone (**Supplementary Figure 1**). All study communications were conducted virtually via video/phone calls, text messages, and emails. Participants assigned to the acupressure arm received training in self-administering acupressure and had access to the training materials saved on a HIPAA-compliant server to facilitate refreshment as needed. Those in the acupressure plus usual care arm completed 6 months of monthly follow-up, whereas those receiving usual care alone were followed monthly for 3 months. The study team maintained routine and real-time communications with each participant to facilitate adherence and remote data collection throughout the participation.

**Table 1.**
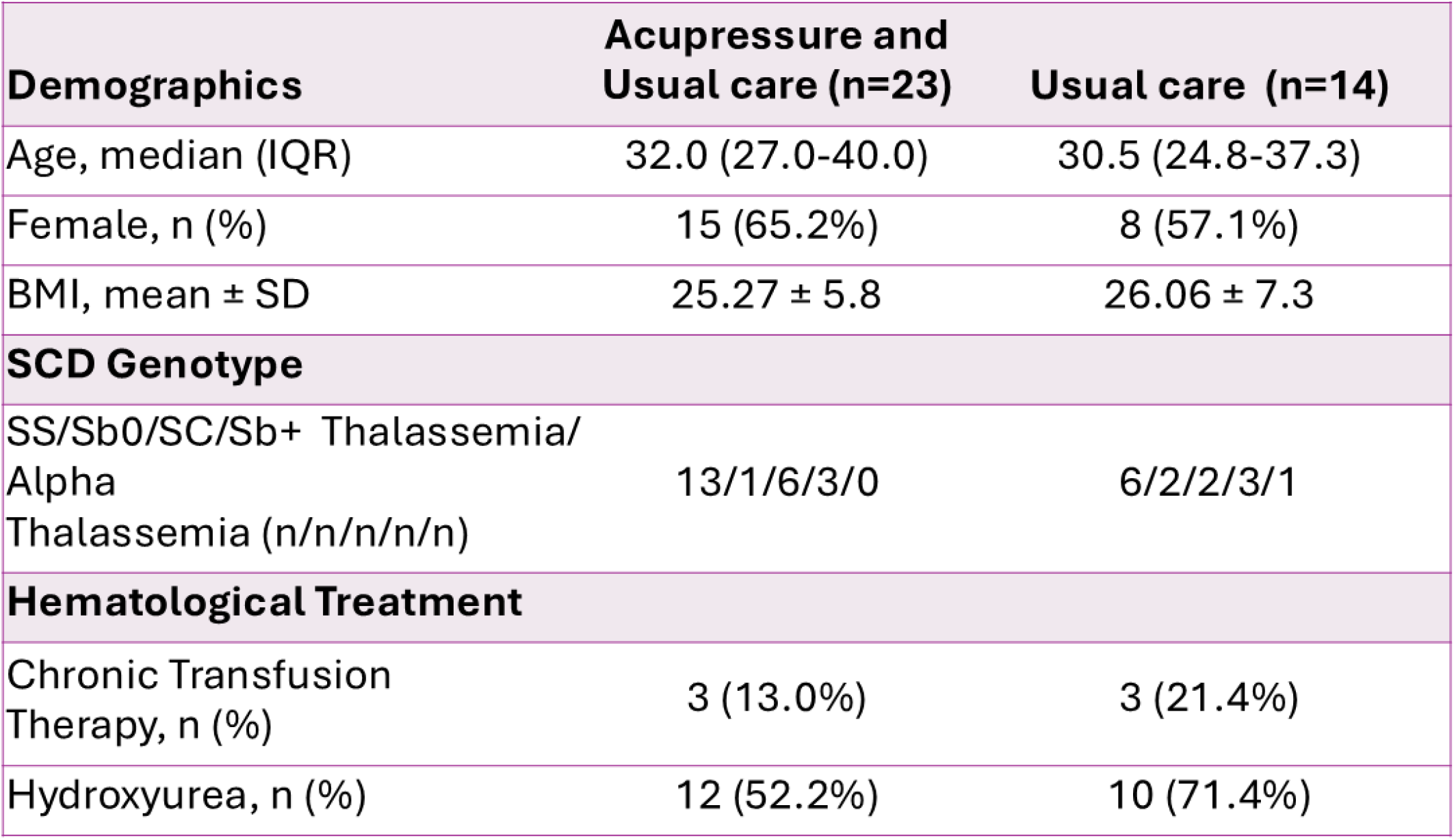
Subjects Demographics.

### Treatment protocol

Participants who were randomized to the acupressure group received training to administer the acupressure treatment every other day for 5 weeks. The acupoints used in this trial (ST36, SP6, SP10, LR3, LI4, LI11, DU24, DU20, Yin Tang) were selected from the main acupoints of our ongoing RCT (ClinicalTrials.gov Identifier: NCT05045820).

### Patient-reported outcome measures (PROMs)

PROMs were collected before, during (weekly), and after the 5-week treatment period to evaluate changes in pain and co-morbidities. The Brief Pain Inventory was used to assess pain severity. Pain interference, anxiety, depression, fatigue, and sleep disturbance were assessed using PROMIS-29. The Widespread Pain Index evaluated the spatial extent of pain by the number of body sites endorsed as painful on a body map. QoL was determined using the Pediatric Quality of Life Inventory (PedsQL, both pediatric and adult versions).

### Statistics

Linear mixed effect models were applied to analyze the treatment’s effectiveness by comparing the PROM results among N=23 acupressure participants and N=14 usual care-only participants as compared to their respective baseline at 1) pre- and post-treatment; and 2) individual timepoints pre-, during, and post-treatment phases. Under the same mixed effects models, PROM results at time points after pre-treatment were further compared to their pre-treatment levels for each group separately. Participant-level random intercepts were included to account for within-participant correlations. Time points were included as a categorical variable for potentially nonlinear temporal trends. Given that VOCs and other major life and/or health events may have severe confounding effects on outcomes, models also evaluated the presence of a stressor at each timepoint (coded as yes or no). Analyses were conducted in SAS 9.4 (SAS Institute, Cary, NC, USA). Two-sided P values <0.05 were considered statistically significant. Since this was a pilot study mainly to generate hypotheses instead of to test hypotheses, no adjustments for multiple comparisons were made.

## 3. Results

Of the 45 participants consented, five were lost to follow-up and one withdrew. The remaining 39 individuals (n=9 non-randomized individuals with treatment plus usual care and n=30 randomized individuals; average retention rate: 82.95%) were active throughout the follow-up phase, and two were excluded from this analysis for poor data quality based on incongruent responses between similar surveys (**Supplementary Figure 1**). There were no significant differences in age, BMI, or sex between the two groups. Among the total of 391 scheduled treatment sessions for the 23 SCD participants receiving acupressure, only 35 treatment sessions were missed (91.0% adherence). The high adherence rates suggest that the treatment regimen was feasible for most participants. No participants in the treatment group reported side effects.

The acupressure group exhibited significant reductions in all measures following the 5-week treatment (**Figure 1**), as compared to the usual care only group including pain severity (mean reduction=−2.60, 95% CI [-3.95, -1.24], p<0.001), pain interference (mean reduction=-3.22, 95% CI [-5.66, -0.78], p=0.011), physical dysfunction (estimate=-2.51, 95% CI [-4.44, -0.58], p=0.012), fatigue (mean reduction=-4.98, 95% CI [-7.83, -2.13], p=0.001), sleep disturbance (mean reduction=-4.02, 95% CI [-6.29, -1.75], p=0.001), anxiety (mean reduction=-2.14, 95% CI [-4.05, -0.23], p=0.029), depression (mean reduction=-3.13, 95% CI [-4.85, -1.40], p=0.001), and widespread pain (mean reduction=−2.98, 95% CI [-4.91, -1.04], p=0.004). QoL also signified improvement in the treatment group (mean improvement=9.43, 95% CI [2.64, 16.21], p=0.008). Stressors significantly impacted the outcomes of pain severity (mean effect=+2.94, p<0.001), pain interference (mean effect=+2.43, p=0.043), and widespread pain (mean effect=+2.46, p=0.015) when present.

**Figure 1.**
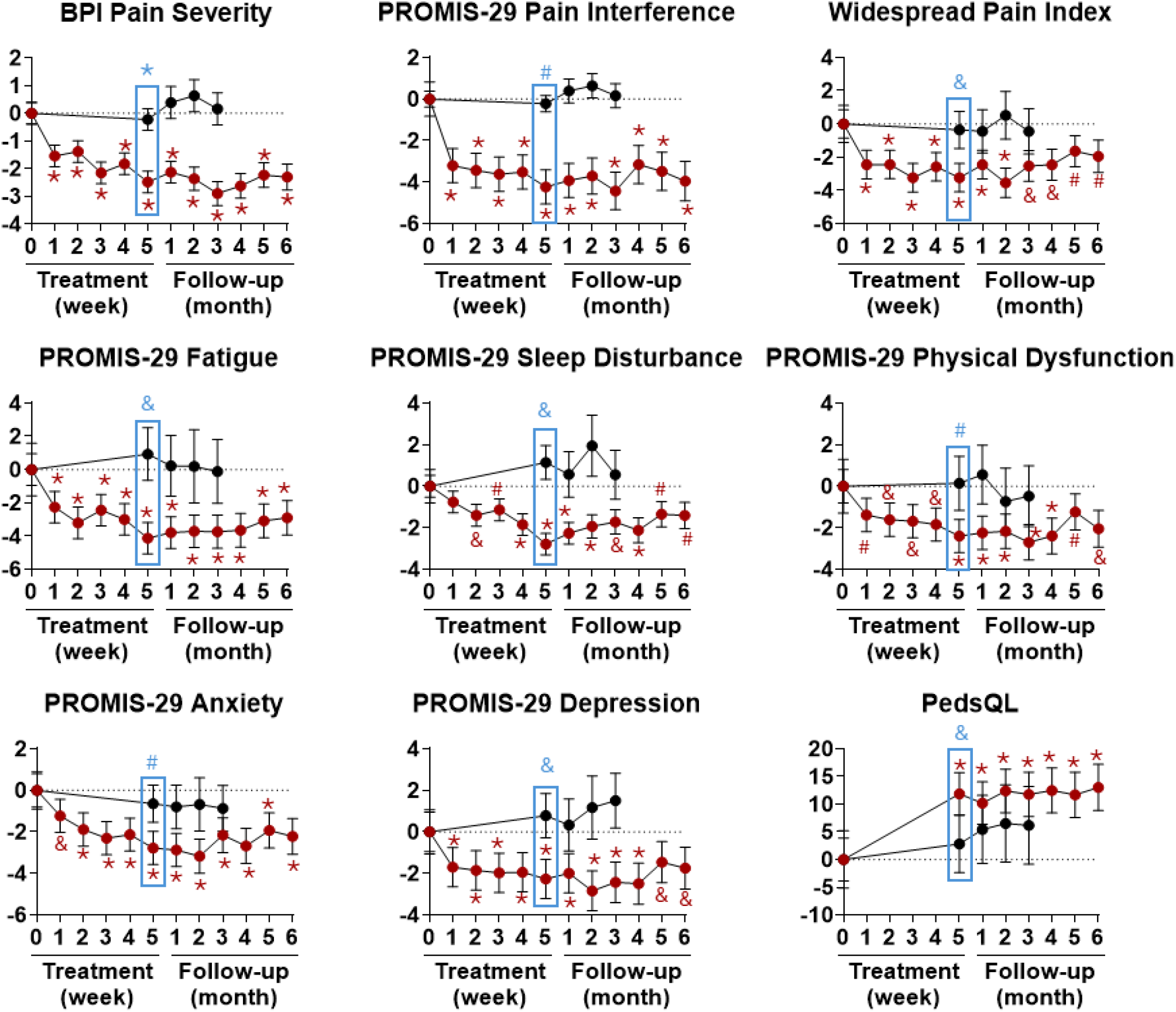
Longitudinal changes in patient-reported outcomes during the 5-week treatment phase and 6-month follow-up visits. Each panel depicts the within-subject mean change from baseline (post treatment minus baseline) for each timepoint. Black symbols represent the usual care only group, while red symbols represent the acupressure plus usual care group. Negative values reflect improvement except in quality of life. Blue boxes represent the group comparison at the 5^th^ week timepoint between the two groups. BPI, Brief Pain Inventory; PedsQL, Pediatric Quality of Life Inventory *p<0.001, ^&^p<0.01, ^#^p<0.05 compared to baseline.

Longitudinally, participants receiving acupressure in addition to usual care demonstrated greater and more sustained improvements compared to their baseline in SCD-related symptoms than those receiving usual care alone (**Figure 1**). Pain severity, pain interference, widespread pain, fatigue, anxiety, and depression declined from baseline to week 5 and remained lower throughout follow-up in the acupressure group. Physical dysfunction improved steadily from week 1 and remained significant throughout the follow-up period. Sleep disturbance did not show significant change until week 2, then continued to improve and remained significant throughout the follow-up period in the acupressure group.

## 4. Discussion

Participants who underwent acupressure reported reduced pain, anxiety, depression, fatigue, sleep disturbances, and physical dysfunction, along with improved QoL. Symptom improvements typically reached their peak by the fifth week of treatment and lasted for at least three months afterward. The treatment was well tolerated, evidenced by high adherence rates, and no side effects were reported. These findings suggest that self-administered acupressure may be an effective treatment and well-suited for telehealth applications in SCD.

This study is limited by a small sample size. Furthermore, the remote self-administered acupressure introduces potential confounding factors associated with the consistency of treatment delivery, as participants must accurately identify acupoints and maintain consistent administration at each session. Although participants were trained and had ongoing access to training materials, potential variations in protocol are a concern for full replication. Large-scale RCT incorporating SCD populations are needed to validate these preliminary findings and further elucidate acupressure’s potential as an effective telehealth approach for SCD pain management.

## Data Availability

Deidentified data is available upon reasonable request from author Y.W.

## Acknowledgement

This research received funding from an NIH K99/R00 award (Grant # 5R00AT010012) and the Indiana University Health – Indiana University School of Medicine Strategic Research Initiative awarded to Y.W.

## Authorship Contributions

L.H. drafted the manuscript and assisted with data collection; A.Q.P. coordinated study procedures, assisted with data collection, and helped revise the manuscript; M.J. assisted with recruitment and data collection of participants recruited at the California site; Z.L. performed the data analysis; Y.W. and A.R.W.O directed patient recruitment at Indiana site; Z.P. directed patient recruitment at the California site and revised the manuscript; S.E.H. and R.E.H. helped revise the manuscript; Y.W. revised the manuscript, developed the conceptual framework, and directed the overall performance and quality of the clinical investigation.

