## Supplementary Figure 1 for "Acupressure alleviates pain and clinical symptoms in patients with sickle cell disease"

**Supplementary Figure 1: CONSORT 2025 Flow Diagram**

Assessed for randomization (n=33)

Non-randomized (n=12)

Consented (n=45)

Analyzed for primary outcome (n=14)

Excluded from analysis (give reasons) (n=1)

Data Quality

Analyzed for primary outcome (n=14)

Excluded from analysis (give reasons) (n=1)

Data Quality

Analysis

Discontinued intervention (give reasons) (n=0)

Lost to follow-up for primary outcome (give reasons) (n=2):

Discontinued control (give reasons) (n=0)

Lost to follow-up for primary outcome (give reasons) (n=1):

Excluded (n=0)

Not meeting inclusion criteria (n=0)

Declined to participate (n=0)

Other reasons (n=0)

Randomized (n=33)

Allocation

Follow-Up

Allocated to intervention (n=17)

Received allocated intervention (n=17)

Did not receive allocated intervention (give reasons) (n=0)

Allocated to control (n=16)

Received allocated intervention (n=16)

Did not receive allocated intervention (give reasons) (n=0)

Enrollment
